# Can 10x cheaper, lower-efficiency particulate air filters and box fans complement High-Efficiency Particulate Air (HEPA) purifiers to help control the COVID-19 pandemic?

**DOI:** 10.1101/2021.12.04.21267300

**Authors:** Devabhaktuni Srikrishna

## Abstract

Public health departments such as CDC and California Department of Public Health (CA-DPH) advise HEPA-purifiers to limit transmission of SARS-CoV-2 indoor spaces. CA-DPH recommends air exchanges per hour (ACH) of 4-6 air for rooms with marginal ventilation and 6-12 in classrooms often necessitating multiple HEPA-purifiers per room, unaffordable in under-resourced community settings. Pressure to seek cheap, rapid air filtration resulted in proliferation of lower-cost, Do-It-Yourself (DIY) air purifiers whose performance is not well characterized compared to HEPA-purifiers. Primary metrics are clean air delivery rate (CADR), noise generated (dBA), and affordability ($$). CADR measurement often requires hard-to-replicate laboratory experiments with generated aerosols. We use simplified, low-cost measurement tools of ambient aerosols enabling scalable evaluation of aerosol filtration efficiencies (0.3 to 10 microns), estimated CADR, and noise generation to compare 3 HEPA-purifiers and 9 DIY purifier designs. DIY purifiers consist of one or two box fans coupled to single MERV 13-16 filters (1”-5” thick) or quad filters in a cube. Accounting for reduced filtration efficiency of MERV 13-16 filters (versus HEPA) at the most penetrating particle size of 0.3 microns, estimated CADR of DIY purifiers using 2” (67%), 4” (66%), and 5” (85%) filters at lowest fan speed was 293 cfm ($35), 322 cfm ($58), and 405 cfm ($120) comparable to best-in-class, low-noise generating HEPA-purifier running at maximum speed with at 282 cfm ($549). Quad filter designs, popularly known Corsi-Rosenthal boxes, achieved gains in estimated CADR below 80% over single filter designs, less than the 100% gain by adding a second DIY purifier. Replacing one of the four filters with a second fan resulted in gains of 125%-150% in estimated CADR. Tested DIY alternatives using lower-efficiency, single filters compare favorably to tested HEPA-purifiers in estimated CADR, noise generated at five to ten times lower cost, enabling cheap, rapid aerosol removal indoors.

## Introduction

Recent studies demonstrate live SARS-CoV-2 in micron^1 2^ and submicron^3^ aerosols from the exhaled breath of infected people. Pathogen-carrying aerosols have potential to accumulate in airspaces of poorly ventilated, indoor spaces such as classrooms, clinics, offices, homes, restaurants, and bars and other community settings, and if inhaled may result in COVID-19 infections.^4^ Separately, toxic aerosol pollution such as from wildfires, wood burning, and other sources can be encountered in these same locations with ventilation using unfiltered, outdoor air. Portable air filtration in the form of commercially available, high efficiency particulate air purifiers (HEPA-purifiers) is useful to remove both types of aerosols without relying on centralized air-handling systems designed to turn on / off to control temperature, not necessarily to run continuously to sanitize the air.

Indoor ventilation upgrades, HEPA-purifiers, and respirators (e.g. N95) can reduce inhalation of SARS-CoV-2 and its variants, other airborne viruses, aerosolized bacteria, and particulate pollution, including allergens such as pollen, dust, bacteria, fungi, etc. However, purifiers/ventilation can be expensive^71^ and respirators can be difficult to wear for an extended period of time causing many organizations or households to hesitate. To mitigate COVID exposures in non-healthcare settings (including businesses, companies, offices, restaurants, schools, faith-based organizations, etc.), recent guidance by California Department of Public Health is that air in rooms with marginal ventilation be filtered 4 to 6 times per hour (i.e. 4-6 air exchanges per hour or ACH), and in classrooms, 6 ACH minimum with as high as 12 ACH preferable (page 5 of “Ventilation and Filtration to Reduce Long-Range Airborne Transmission of COVID-19 and Other Respiratory Infections: Considerations for Reopened Schools”^90^). A recent study from Italy suggests infection rates were cut by 40% to 80% in classrooms that implemented mechanical ventilation of from 2 to 6 ACH respectively still allowing some level of transmission.^84 87^ For comparison, CDC and WHO recommend an airborne isolation room in a hospital to have a minimum of 12 ACH based on the time required for airborne-contaminant removal^54^ and estimated risk of infection.^91^ To meet or exceed any of these ACH targets, high price-points for select HEPA-purifier models without incurring excessive noise generation make them unaffordable for many households and communities.

For example, in a typical classroom (30’x30’x8’), a HEPA-purifier with clean air delivery rate (CADR) of 300 cubic feet per minute (cfm) needs 24 minutes to cycle the air once (2.5 air exchanges per hour or ACH) under ideal mixing conditions.^5 6^ This classroom would require 2-3 air HEPA-purifiers with low-noise generation costing $400-$500 each (with periodic $40-$80 filter replacements) to achieve 4-6 ACH costing approximately $1000-$1500 per classroom, outside the budget of many schools. If needed, getting to 12 ACH as in hospitals would require proportionally more HEPA-purifiers putting it further out of reach.

Lower-cost alternatives to HEPA-purifiers have been in use for a decade, but their air filtration and safety properties have not been examined in detail until recently. A Minimum Efficiency Rating Value-13 (MERV-13) filter attached to a box fan (e.g. using duct tape, bungee cord, rubber band, clips) was demonstrated to effectively filter wildfire PM2.5 and submicron particles.^7^ Ford Motor Company sponsored “Scrappy Filtration” which donated 20,000 box-fan and air filters held together with a cardboard structure for use in classrooms to underserved communities.^8 9^ However, the published literature regarding these lower-cost box-fan-filters is lacking key metrics such as filtration efficiency (%), air flow (CADR), and noise generation (dBA) and does not use lower-cost, rapid test methods (e.g. that do not require a potentially more expensive, sealed test chamber) which would more readily enable comparisons to a wider range of HEPA-purifiers.

Below, we compare cost-effectiveness of aerosol filtration by best-in-class, portable HEPA-purifiers and do-it-yourself (DIY) alternatives built from box fans and HVAC filters rated at MERV-13, 14, 15 and 16. Comparison is based on three metrics: estimated clean air delivery rate (CADR), the noise generated (dBA), and initial cost or affordability ($). Measurement devices include an ISO-certified, calibrated aerosol meter for input/output particle filtration of sizes ranging from 0.3 μm to 10 μm, airspeed meter to measure airflow, and NIOSH sound meter app to measure noise. Leveraging these test methods, we were able to evaluate and compare three different HEPA-purifiers against common DIY fan-filter configurations.

Tested DIY configurations include box fans with six single-filter configurations in Table 2, and five multiple filter configurations in Table 3 (four quad filter with one box fan and one triple filter with two box fans). Each configuration utilized the same make/model of box fan tested at speeds 1 and 3 as shown in the columns of Tables 2 and 3. Specifications are provided in the Methods section. Tested configurations with the quad filter design are based on a cube formation, popularly known on social media as the Corsi-Rosenthal box, proposed to enhance airflow and CADR over single filter designs. In order to examine how CADR is impacted by additional flow, the Corsi-Rosenthal DIY cube (4 filters and 1 fan) was compared to a cube design replacing one of the four filters with an additional fan (i.e., 3 filters 2 fans). A preview of these results showing estimated CADR versus cost is shown in Figure 1 with examination of the multidimensional tradeoffs between filtration efficiency at different particle sizes, airflow, estimated CADR, noise generated, number of fans and filters, and unit cost in the remainder.

**Figure 1:**
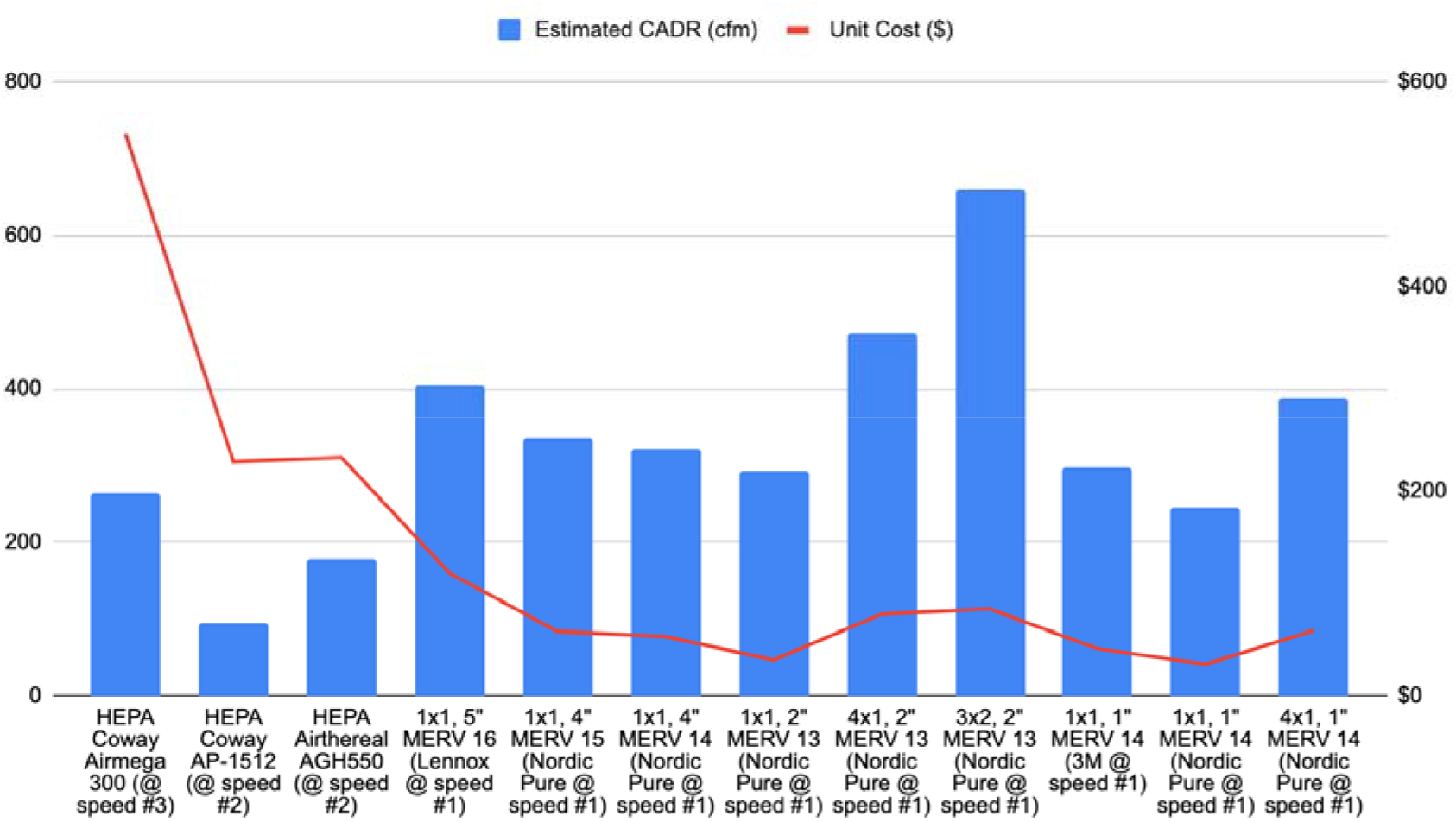
comparison of CADR (estimated by airflow x filtration efficiency) and cost of HEPA purifiers and DIY box fan-filters e.g., 3×2 means three filters were used with two box fans.

### Safety Notes

California Air Resource Board recommends never leaving box-fan air filters unattended while turned on, and to use box fans manufactured after 2012 and are clearly identified with the UL or ETL safety markings because it is likely they have a fused plug to prevent electrical fires e.g. if the device is inadvertently knocked over.^10^ Chemical Insights, a subsidiary of Underwriters Laboratories, recently tested five different electric box fan models (approximately 20” × 20” in size) with attached air filters and concluded that all measured temperatures fell below the maximum acceptable thresholds defined by the market safety standard for electric fans (UL 507).^11^ In detail, in addition to assessing DIY fans with clean MERV 13s, DIYs were tested with filters loaded with two types of particles (ASHRAE dust and smoke from biomass burning to represent a loaded filter due to a wildfire), and test scenarios also included fully sealing the box fans in plastic and running fans for over 7 hours face-down on the floor. Even under those scenarios, the fans’ surface components did not reach temperatures that would cause minor burn/injury, and all fan models were able to operate continuously throughout all test scenarios without reaching UL 507 thresholds. The resources and information in this article (the “Content”) are for informational purposes only and should not be construed as professional advice. The Content is intended to complement, not substitute, the advice of your doctor. You should seek independent professional advice from a person who is licensed and/or qualified in the applicable area. No action should be taken based upon any information contained in this article. Use of the article is at your own risk. Patient Knowhow, Inc. takes no responsibility and assumes no liability for any Content made available in this article.

### Usage Notes

Commercially available HEPA-purifiers typically have “smart” settings. Based on the presence of particulate matter in the air, they use a built-in sensor to automatically adjust their fan speed. These built-in sensors are not typically triggered by the respiratory aerosols that carry SARS-Cov-2 viruses. Although this maybe acceptable if you’re concerned only about smoke, if the goal is to reduce COVID-19 risk, fan speed settings for HEPA-purifiers need to be set to operate at a constant speed to continuously filter the air regardless of particular matter detected by the sensor, higher the better. A best practice for shared spaces such as classrooms and offices where viral transmission is a primary concern is to run the HEPA-purifier at its maximum fan speed or the highest fan speed tolerable to the occupants of the room.

## Methods

Traditional methods that measure the effect of the air purifier (and its CADR) using the decay rate of aerosols generated or injected into an isolated, unoccupied room require greater investment of time, effort, risk of inhalation of the generated aerosols, and training/expertise compared to the much simpler DIY test methods described here.^78 79^ We use ambient aerosols to rapidly compare different filters, fans, and configurations with low-cost, measurement tools available online (e.g. Amazon) also making these results easy to replicate and verify with limited resources.

### HEPA-purifiers

Three models evaluated include the Coway Airmega 300 ($549, 285 cfm), Coway AP-1512HH ($229, 233 cfm), and Airthereal AHG550 ($233, 324 cfm). Unit prices are as listed on Amazon and “smoke” clean air delivery rate (CADR) in units of cubic feet per minute (cfm) is as listed on manufacturer website. The HEPA air purifiers were chosen to be a selection of models known to be most quiet or well-reviewed. Note: “smoke” in this context of manufacturer-reported CADR typically refers to tobacco smoke not wildfire smoke. The tests for the HEPA air purifiers were conducted on November 11, 2021.

### HVAC filters with box fan

For DIY configurations, the box fans used are 20” Lasko Fan ($20) along with 20” x 20” HVAC filters made by Nordic Pure of width and rating 1” MERV-14 ($11), 2” MERV-13 ($15), 4” MERV-14 ($38), 4” MERV-15 ($42), by 3M of width and rating 1” MERV 14 ($26), and by Lennox of width and rating 5” MERV-16 ($99). MERV stands for minimum efficiency reporting values. The single and quad filter designs pictured in Figure 2 use one fan, and the triple filter design uses two fans with one fan replacing one of the filters. The square-shaped 20” box fans tend to pull in air from their front at their corners of the box^80 81^, and a cardboard shroud of 14” diameter was attached to the output side (front) of each fan to enhance airflow through the filter by reducing flow recirculation (via the corners of the front of the box fan) and eddies at the fan blade tips. For comparison, three methods of attaching the box fan to the air filters included vacuum (i.e. no attachment just relying on airflow), duct tape, and velcro. Unit prices are as listed on Home Depot, Amazon, or Walmart. The tests for the HVAC filters with box fans (DIY air purifiers) were conducted January 10 to 13, 2022.

**Figure 2:**
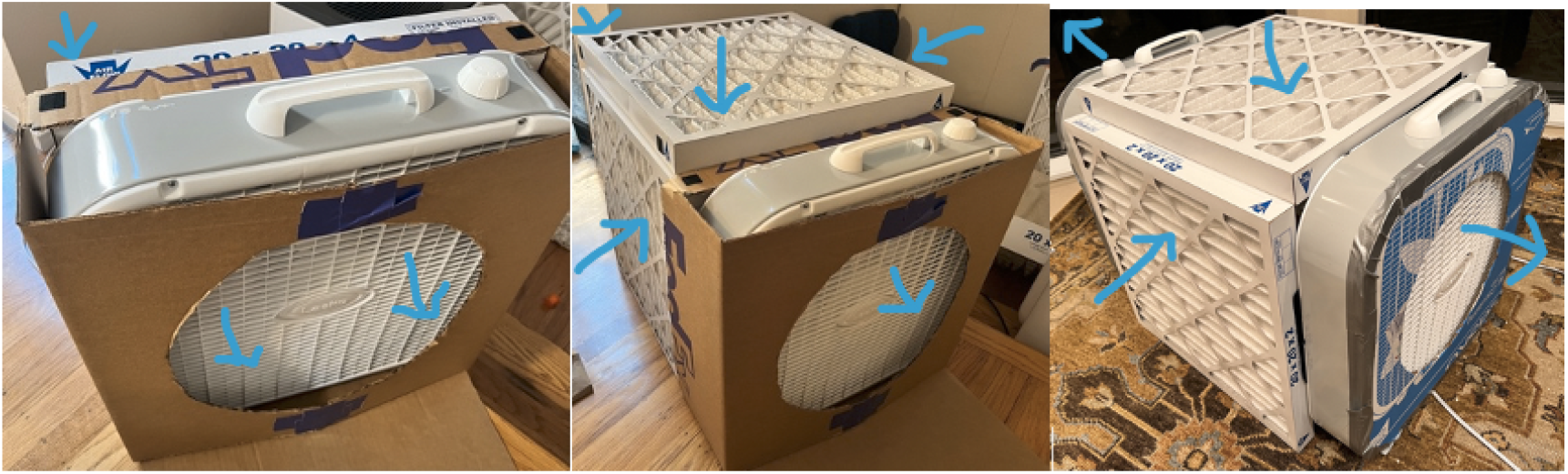
Do-It-Yourself Box Fan and Filters (single, quad filter designs with one fan and triple filter design with two fans). Blue arrows indicate direction of airflow.

### Aerosol filtration efficiency

For each HEPA-purifier or box fan filter, counts at each particle size bin (0.3, 0.5, 0.7, 1.0, 2.5, 5, and 10 μm) were tested using an ISO-certified, particle counter (Temtop Particle Counter PMD 331 available on Amazon) with the each device (its fan) turned ‘on’ and ‘off’ consecutively. The tests were conducted in a house near an open window with fan blowing air through the window to allow for aerosols (particles) from outdoors to continuously enter the room. Five counts were recorded and averaged on the particle counter when its input was placed directly at the output of the air purifier/filter for 30 seconds. The filtration efficiency^89^ at each particle size was estimated by the formula, efficiency = 1 - on / off. To compare each fan-filter configuration in this study, the filtration efficiency depends only on the ratio of on to off, not on their absolute values. The five counts were recorded individually over no more than 15 minutes, however the filtration efficiencies were computed using consecutively recorded counts no more than 2 minutes apart.

### Air flow

For each HEPA-purifier or DIY box fan filter, the airspeed was measured using an anemometer (BTMETER BT-100 Handheld Anemometer available on Amazon) held at the output perpendicular to the airflow. An average of three or more airspeed measurements (feet per minute) were multiplied by area of output to estimate the airflow (cubic feet per minute).

### Noise

Noise measurements were taken for each air purifier/filter using an iPhone app maintained by National Institute for Occupational Safety and Health (NIOSH).^12^ Three or more noise measurements were taken at a 9” distance perpendicular to the direction of the output airflow and averaged.

### CADR (estimated)

For each HEPA-purifier or DIY box fan filter, the Clean Air Delivery Rate (CADR) was estimated as the product of the measured air flow (cfm) and aerosol filtration efficiency (%) at 0.3 microns.

## Results

Note: Measurements in each table below are shown as averages (standard deviation in parentheses).

### HEPA-purifiers

As shown in Table 1, the results for the HEPA-purifiers show particle filtration efficiencies near or exceeding 95% at most particle sizes ranging from 0.3 to 10 μm and higher in many cases. At their maximum fan speed, the noise generated by the Coway Airmega 300 (59.3 ± 2.1 dBA) was roughly ∼5 dBA lower than the the Coway AP 1512 (64.7 ± 0.57 dBA) and Airthereal (66.3 ± 0.6 dBA). To achieve comparable noise levels to the Coway Airmega 300, both the Coway AP-1512 and the Airthereal AGH550 had to be set at fan speed 2 which cut their estimated airflow (CADR) roughly in half. As a verification of the estimation methodology, the estimated CADR using airflow (cfm) and filtration efficiency at 0.3 microns was within 10% of the the smoke CADR rating from the manufacturer’s website. Based on this data it also appears some HEPA air purifiers may have higher or lower rate of filtration at 0.3 microns than others although well below the 99.97% typically assumed for HEPA. These differences appear to reflect properties of the filters themselves rather than sample size. In Table 1, the standard deviations for the filtration efficiency at 0.3 microns are small even for five measurements (3.5% for Airthereal and less than 1% for both of the Coway models). This suggests it is not simply a reflection of the sample size of 5 measurements but rather a property of the filters themselves. There is even an observable difference between the two Coway models as shown in Table 1.

**Table 1:** Performance of HEPA-purifiers.

| Setup | | Noise and Airflow | | | CADR (cfm) | | Filtration Efficiency (%) avg of five measurements at each particle size bin ($\mu\text{m}$) | | | | | | |
| --- | --- | --- | --- | --- | --- | --- | --- | --- | --- | --- | --- | --- | --- |
| | Price (est.) | Fan speed | Noise (dBA) avg. of three samples 9" away | Est. Airflow (output) cfm | From Manufacturer (smoke CADR) | Est. using airflow x filtration eff. @ 0.3 $\mu\text{m}$ | 0.3 | 0.5 | 0.7 | 1.0 | 2.5 | 5.0 | 10.0 |
| Coway Airmega 300 | \$549 | 3 | 59.3 (2.1) | 282 (61) | 285 | 265 | 94.1 (0.9) | 94.5 (1.1) | 97.0 (0.96) | 97.4 (0.92) | 98.3 (1.5) | 98.3 (1.8) | 98.0 (1.6) |
| Coway AP-1512 | \$229 | 3 | 64.7 (0.57) | 223 (3.7) | 233 | 216 | 97.4 (0.3) | 98.1 (0.4) | 98.5 (0.6) | 98.7 (0.8) | 98.4 (1.7) | 98.0 (3.0) | 97.6 (5.5) |
| Coway AP-1512 | \$229 | 2 | 46.0 (3.5) | 97 (5.7) | | 94 | | | | | | | |
| Airthereal AGH550 | \$233 | 5 | 66.3 (0.6) | 410 (81) | 324 | 354 | 86.5 (3.5) | 89.5 (0.9) | 90.6 (2.1) | 92.7 (1.9) | 93.9 (3.2) | 96.0 (2.4) | 98.6 (2.1) |
| Airthereal AGH550 | \$233 | 2 | 47.3 (1.5) | 206 (42) | | 178 | | | | | | | |

### Single filter configurations with box fan

The noise generated by the box fan was tolerable (subjectively to the author, without it feeling stressful) only at fan speed #1 (out of three) with a measured noise level of 54 dBA comparable to the Coway Airmega 300 at 59 dBA. As shown in Table 2, the filtration efficiency of the MERV-13, 14, 15, or 16 filters at the most penetrating particle size of 0.3 μm varied by thickness from approximately 60% to 85% with highest observed for MERV-16 filter and lowest for the MERV 13 filter, well below the HEPA filters. Even though MERV-13 filters may remove less than 80% of particles in just one pass, if the air recirculates multiple times through filter the aerosol concentrations can be rapidly reduced.^7^ The filtration efficiency exceeded 90% in some cases at particle sizes above 1 μm.

**Table 2:**
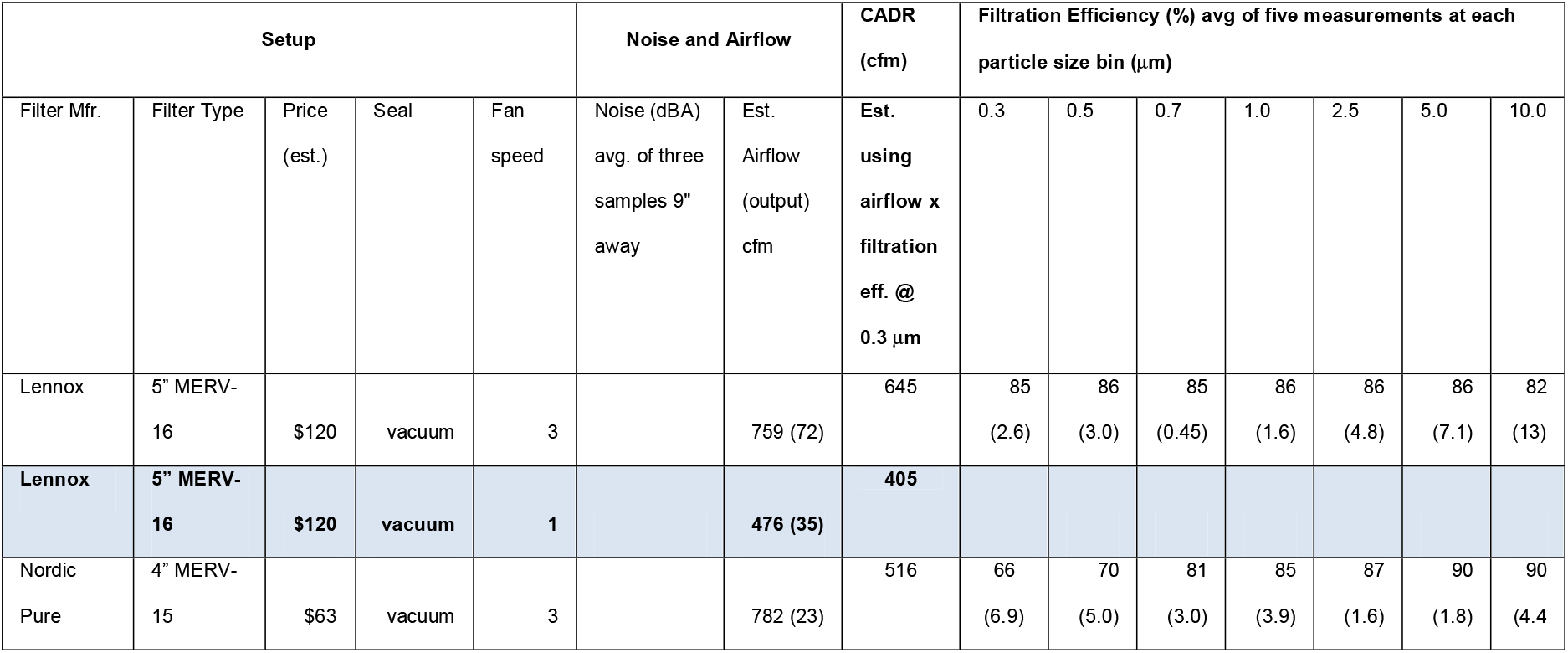
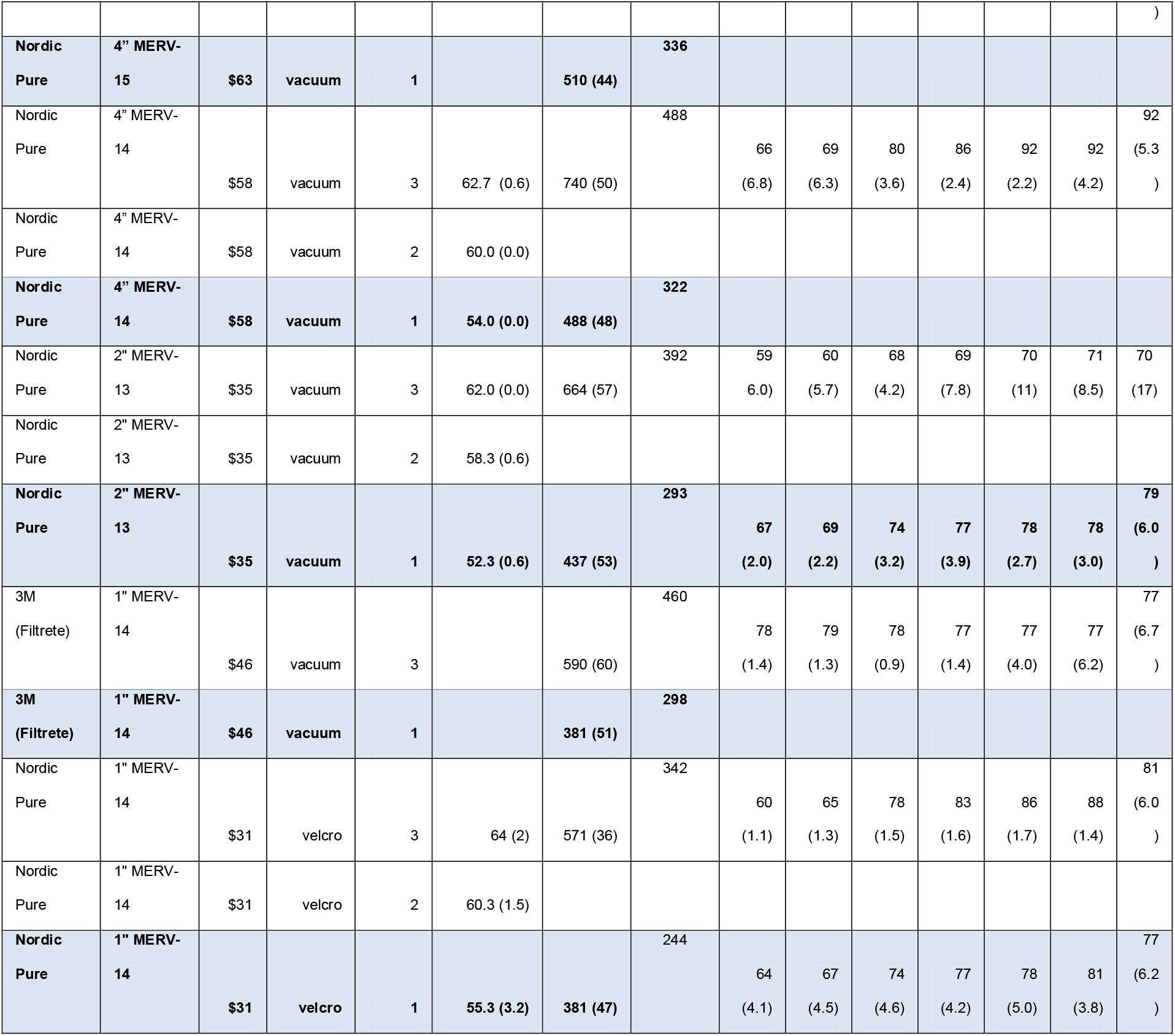
Performance of single-filter configurations with a box fan.

For all filters, filtration efficiencies were measured at fan speed #3, but also at fan speed #1 for the 2” MERV 13 and 1” MERV 14 filters at which the filtration efficiencies were higher. In general, that is expected to hold true with the other filters so the estimated CADR for each filter at fan speed #1 (above) would be commensurately higher as well.

### Cube configurations with box fan(s)

Using comparably rated filters and fan speeds, the quad filter design with one box fan (4×1), popularly known as the Corsi-Rosenthal box, showed significant improvement in airflow and filtration efficiency over single filters (Table 3). Whereas results from attaching the filters were similar with duct tape (to seal up any gaps) or velcro (leaving small airgaps), suggesting any leaks created by a loose fit may not make much difference. Notably, the filtration efficiency of the quad filter design was marginally better than the corresponding single filter design possibly due to lower pressure across the filters. The three extra filters in the CR box (4×1) increased airflow by less than 50% over equivalently rated single filter configuration (1×1), whereas it took adding an extra fan and two extra filters in the cube formation (3×2) to double the airflow and estimated CADR.

**Table 3:**
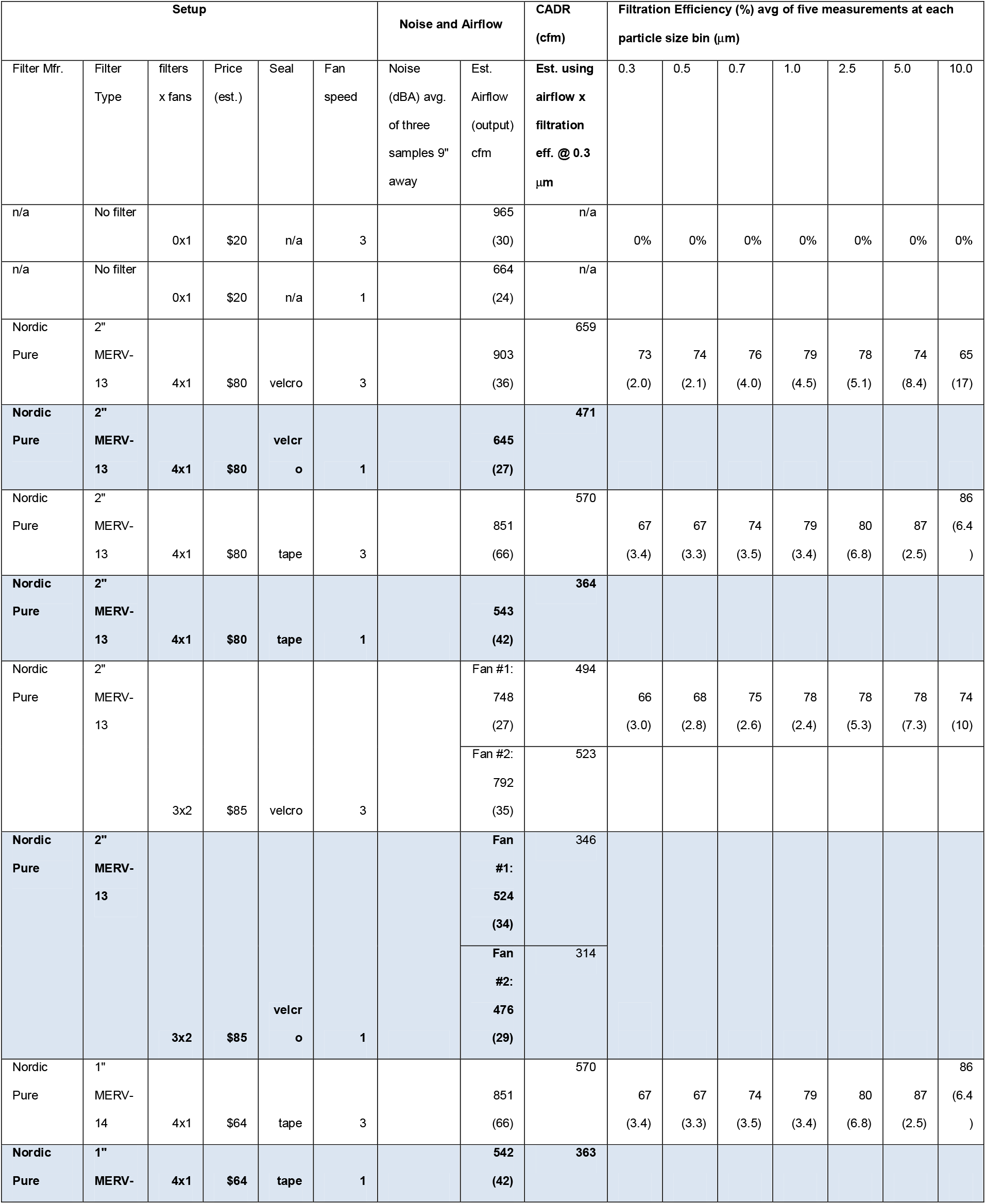
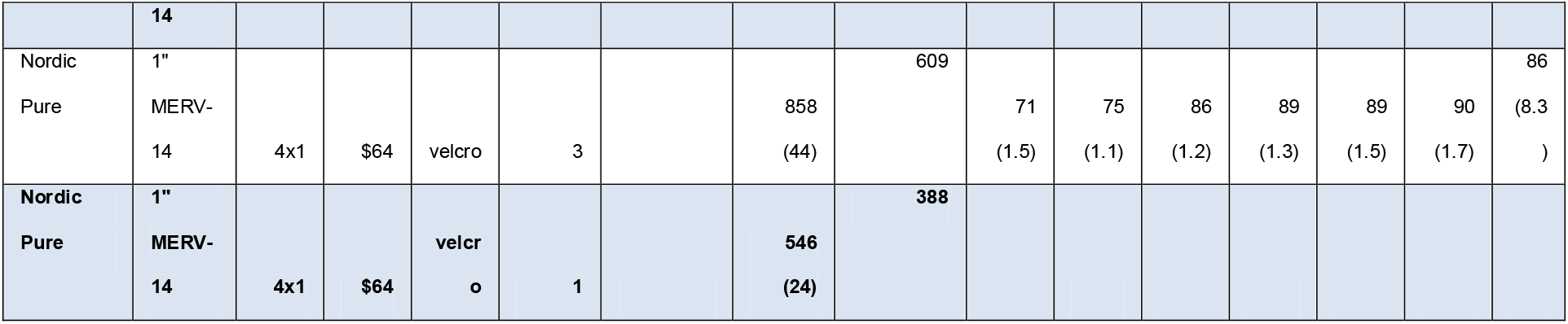
Performance of cube configurations (filters x fans: 4×1 and 3×2)

## Discussion

Accounting for lower filtration efficiency of MERV-13,14,16 filters at the most penetrating particle size of 0.3 μm compared to HEPA, the estimated clean air delivery rate (CADR) of a do-it-yourself (DIY) setup using 2” MERV-13 (67%), 4” MERV-14 (66%), 5” MERV 16 (85%) filters with a box fan running at fan speed 1 for tolerable noise was 293 cfm, 322 cfm, 405 cfm as shown in Figure 3. This is comparable or better than a best-in-class, low-noise generating HEPA-purifier running at maximum speed with at 282 cfm. Yet the upfront cost of the components of the DIY setup were approximately 5 to 10 times less ($35-$58-$120) than best-in-class HEPA-purifiers ($549).

**Figure 3:**
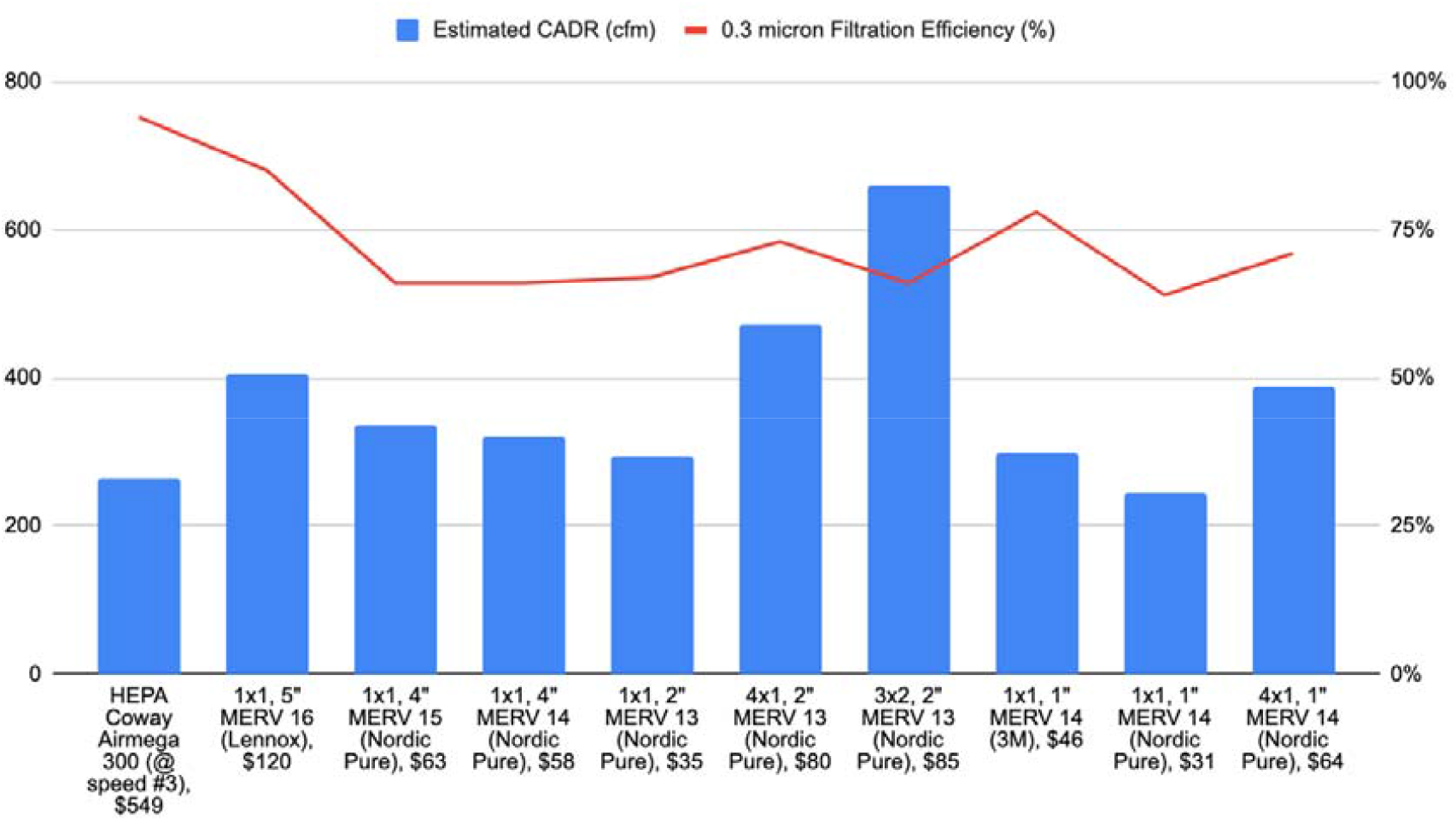
Estimated CADR (cfm) and filtration efficiency (%) for DIY Air Purifiers (filters x fans) @ speed #1 e.g., 3×2 means three filters were used with 2 box fans.

At least two studies have measured the CADR of cube configurations (popularly known as the Corsi-Rosenthal box or CR box) by observing the decay rates of generated aerosols. The first study is by UC-Davis^78^ measured 600 cfm at lowest fan speed to 850 cfm at highest fan speed. The second study is by Illinois Institute of Technology^79^ (IIT), measured below 300 cfm in the range of 0.5 to 3 μm (dust) at the highest fan speed but above 450 cfm in the range of 5-11 μm (pollen). Both used five 2” MERV-13 filters (instead of four in our study) of similar dimensions so it is unclear why the studies by UC-Davis and IIT differ by a factor of two. Our results were in between these two studies but more consistent with the UC-Davis study. The estimated CADR of cube configurations with four 2” MERV 13 filters was 471 cfm at lowest fan speed 1 and 659 cfm at the highest fan speed 3 using 73% filtration efficiency at 0.3 μm (most penetrating particle size). It is possible that greater filtration efficiencies at larger particle sizes may account for this difference with UC-Davis results which is based on generated salt aerosols. Other differences that may account for it are the number of filters (4 vs 5), fans, differences in estimation methodology, box fans from different manufacturers may differ substantially in terms of their flowrate at “free flow” (i.e. without any pressure drops from filters) and when installed in DIY air cleaners with filters, and differences in particles generated and measured at different aerodynamic particle sizes. Further investigation would be required to fully understand why there is such a range.

However at all fan speeds the quad filter design, CR box, did not achieve gains in CADR of more than 80% over a single filter design which is in contrast to the 200-250% gains also reported by UC-Davis (also with shroud and 20” Lasko box fan),^13^ separate from the UC-Davis study in the prior paragraph. CR boxes are unlikely to exceed the airflow with no filters. At fan speed 1, the airflow of quad 1” MERV 14 filter design (546 cfm) was within 25% of that with no filters (646 cfm), and the airflow of the single 1” MERV 14 (381 cfm) was within 50% of no filters. This suggests the quad 1” design is close to optimal but cannot be improved by a factor of 200% over the single 1” filter. At fan speed 1, the airflow with no filter (646 cfm) was measured to be 70% more than the airflow with a single 1” filter (381 cfm), 47% more than 2” MERV-13 filter (437 cfm), and only 32% more than the 4” MERV-14 filter (488 cfm). The cost and extra setup effort from the quad filters (CR box) can be alternatively expended on more simply replicating a new box fan filter (+ 100% gain) so that the airflow is then distributed to different locations in the room or classroom. To illustrate this, replacing one of the 2” MERV-13 filters in the quad filter configuration with a second fan resulted in gains of approximately 125%-150% in estimated CADR (at fan speeds 1 and 3) which is notably better than 100% possibly due to improved filtration efficiency.

Regardless, our results suggest a sweet spot between speed of setup, simplicity, size, and cost is a single 20” box fan with 2” MERV-13 ($35), 4” MERV-14 filter ($58) of 20” length and width. Single filter is easier and simpler to setup than multi-filter designs which is an advantage for scaling up DIY approaches. A second box fan and single filter provide more air cleaning than one CR box with the possibility of benefits for air cleaner placement. Alternatively, if higher filtration efficiency is desired a 5” MERV-16 filter can be used ($120). Given the simplicity and low-cost of the box-fan design using single 2” or 4” MERV-13/14 filters, form-factor improvements at a marginal cost may include a more stable, cardboard box frame so it is not so easy to knock over with printed designs to make it aesthetically pleasing. An aluminum screen fixed to the shroud in front of the fan can help keep small objects (e.g. fingers or pencils) from contacting the fan blade which may be especially important for classroom^70^ and home applications when young children are present.

### Limitations

Although SARS-Cov-2 is extremely small (approximately 100 nanometers in diameter^87^), it may be exhaled in respiratory aerosols and droplets that are much larger (several hundreds of nanometers to several microns). The research on which are the most common COVID transmission modes (aerosol, droplet, or surfaces), and what is the most common aerosol particle sizes by which it is transmitted continues to evolve as new variants emerge. Multiple studies demonstrate SARS-CoV-2 in micron and submicron aerosols from the exhaled breath of infected people.^1 2 3^ In one study, more of the viable virus from human subjects was observed in submicron respiratory aerosols than micron and above.^3^ One potential limitation is in extrapolating results from the ambient test aerosols used in this study to real-world applications for removal aerosols relevant to SARS-Cov-2.

‘Filtration efficiency and CADR results from DIY air cleaners highlight potential performance differences between filters made by different manufacturers of comparably rated MERV 13, 14, 15, 16 filters. Hence filter selection is critical to achieving the results desired. There may also be variability in filter performance due to manufacturing defects or variations among fans and filters from the same manufacturer. The long-term durability of filtration efficiency and airflow of these filters after extended use with box fans is also uncharacterized unlike HEPA-purifiers which have well-understood operational history.

Since measurements of particle concentration were made by placing the inlet of the particle counter towards the outlet of the air purifier, there could be some re-entrainment in the shearing flow leaving the air cleaners that differs as a function of the design of each air cleaner which may impact the measurements.

The aerosol measurements were conducted with a device which is an optical particle counter (OPC) called the PMD 311 manufactured by Temtop. The OPC counts particles in 7 bins (0.3, 0.5, 0.7, 1.0, 2.5, 5, and 10 microns). Possible sources of uncertainty in an OPC based are extremes of relative humidity and differences between optical properties of the aerosol particles measured in this study versus those particles used to calibrate the sensor by the manufacturer.^82^ Extreme relative humidity appears unlikely to be a source uncertainty based on the environmental conditions under which this study was conducted. However, the manufacturer used polystyrene aerosols to calibrate the sensor of the measurement device within 10% at each particle size. The choice of polystyrene material may have some systematic impact on the accuracy of the absolute counts when measuring aerosols made of other materials. The principle of operation of the OPC is it uses a sensor which measures laser light scattered from each particle to estimate its size, with larger (smaller) particles having a greater (lower) scattering cross section which is used to differentiate the size of each particle counted. As shown in Figure 3 of Hagan et, al.,^82^ somewhat surprisingly the scattering cross section of black carbon is higher (or lower) than polystyrene depending on whether particle size is smaller (or larger) than a threshold which is approximately 0.3 microns. If some fraction of the particles that are part of the ambient aerosols measured in our study were much more (or much less) absorptive than polystyrene in a particle-size dependent manner, such as black carbon, it is possible these particles could have a lower (or higher) scattering cross section than the calibration aerosol of same size, respectively. If so they could be counted as part of smaller (or larger) bins than their actual size. The measurements of the OPC in each bin will vary to some extent based on the distribution of absorptive and scattering properties of ambient aerosols relative to the polystyrene aerosol used to calibrate the OPC. However, any such systematic differences between the ambient aerosols and the calibration aerosols might be expected to apply uniformly to both the ‘in’ and ‘out’ measurements and therefore would not necessarily affect the computed filtration efficiency for the particles actually counted (measured) in each bin by the OPC, just that their size distribution might be shifted over by one or more bins based on how each particle’s individual optical properties differs from the calibration aerosol.

For each HEPA-purifier or DIY box fan filter, the Clean Air Delivery Rate (CADR) was estimated as the product of the measured air flow (cfm) and aerosol filtration efficiency (%) at 0.3 microns. Strictly speaking, the CADR estimate would apply to the particles that are counted by the measurement device in the 0.3 micron bin of the Temtop PMD 331 subject to the differences described between calibration and ambient aerosols in the prior paragraph, and therefore may not apply for other particle sizes. The three tested HEPA purifiers are not representative of all HEPA purifiers and likewise tested DIY configurations are not representative of all DIY configurations in terms of cost, noise generation, CADR, etc..

## Conclusions

Given the fact that the SARS-CoV-2 pandemic is largely sustained by superspreading events, there are opportunities to use air cleaning principles to improve the indoor air environments where these superspreading events occur and if applied widely enough to potentially control the pandemic. To date there is no large-scale, real-world data to validate a minimum threshold of ACH to control SARS-Cov-2 transmission in a room. The SARS-CoV-2 pandemic is sustained largely by superspreading events: approximately 80% of cases are transmitted by only 10% to 20% of the infected (super spreaders).^1 14 15^ Unlike healthcare facilities where there are minimum specifications^54^ for ventilation and air filtration in many cases exceeding 15 air exchange per hour (ACH), in community settings^55^ and schools^56^ the CDC (US) recommends increasing air filtration “as high as possible.” Specific targets for ACH in community settings are needed.

Best practices recommended by the state of California in the US are only 4 to 6 air exchanges per hour (ACH),^58^ and 6 to 12 ACH in classrooms.^90^ As an illustrative example, the University of San Diego publicly posts its ACH levels for classrooms, with almost all classrooms exceeding 6 ACH and many exceeding 12 ACH.^93^ A UK study^85^ in a hospital setting measured reduced viral load of indoor air in rooms with Covid positive subjects when air is filtered less than 4.5 ACH versus faster than 9 ACH (Figure 5c of that study). Another study^92^ in a hospital recorded (super) spreading of SARS-CoV-2 in general wards with 6 ACH but also noted the virus was undetectable in the air of airborne isolation rooms at 12 ACH (See the Discussion section of that study). A recent study from Italy suggests infection rates were cut by 80% in classrooms that implemented mechanical ventilation of 6 ACH, suggesting perhaps even 6 ACH may be insufficient to control the spread.^84 87^ For comparison, CDC recommends an airborne isolation room in a hospital to have a minimum of 12 ACH.^54^

Fundamentally, the rate of aerosol (particulate) removal by air filters (HEPA, DIY, etc.) in a room as measured by clean air delivery rate (CADR) must exceed the rate of introduction of these aerosols by a significant margin in order to avoid accumulation of aerosols and achieve rapid clearance (e.g. when introduced by an infected person or smoky source). Outdoor ventilation is generally complementary or beneficial to indoor air filtration (in terms of ACH) except on days when the outdoor air is polluted with wildfire or smoky particulates (e.g. PM 2.5) in which case outdoor air ventilation would introduce additional aerosol further increasing the need for indoor air filtration. Experts at Boston School of Public Health^57^ and the California Department of Public Health^58^ recommend 4 to 6 ACH in community settings to prevent COVID-19. Researchers measuring traces of the SARS-CoV2 coronavirus in the air of a COVID-19 ward of a UK hospital reported they were undetectable once air filtration machines filtered the air in each room every 6 to 12 minutes (5-10 ACH).^73^ Although very effective at removing the virus from the air, achieving these higher ACH targets often incur noise generation intolerable to end-users or may be unaffordable or both. Cost and excessive noise are key factors that limit the use of portable air cleaners.^59 60^

Indoor air cleaning and filtration can also be potentially useful mitigation (to reduce inhalation dose by some amount) in response to other airborne viruses (e.g. respiratory RNA viruses^76^) or release of bioterrorism or biowarfare agents such as anthrax^77 86^ However, these can be unaffordable to much of the US and world population.

As we discovered in the test results, lower-efficiency air filtration by combining off-the-shelf components (box fans with heating, ventilation and cooling or HVAC filters) in tested DIY configurations compares favorably in performance (clean air delivery rate, noise) to the tested HEPA air purifiers but at approximately five to ten times lower cost, and can be an affordable, complementary option for rapid aerosol removal indoors in homes, clinics, schools, offices, and other public venues.

## Data Availability

All data produced in the present study are available upon reasonable request to the authors.

